# Clinico-epidemiological characteristics of asymptomatic and symptomatic COVID-19-positive patients in Bangladesh

**DOI:** 10.1101/2020.08.18.20177089

**Authors:** Mohammad Jahid Hasan, Sayeda Mukta Chowdhury, Md. Abdullah Saeed Khan, Monjur Rahaman, Jannatul Fardous, Tanjir Adit, Mustafizur Rahman, Md. Tarek Hossain, Shakila Yesmin, Enayetur Raheem, Mohammad Robed Amin

## Abstract

**Background:** As of August 15, 2020, Bangladesh lost 3591 lives since the first Coronavirus disease 2019 (COVID-19) case announced on March 8. The objective of the study was to report the clinical manifestation of both symptomatic and asymptomatic COVID-19-positive patients.

**Methods:** A online-based cross-sectional survey was conducted for initial recruitment of participants with subsequent telephone interview by the three trained physicians in 237 adults with confirmed COVID-19 infection in Bangladesh. The study period was between 27 April to 26^th^ May, 2020. Consent was ensured before commencing the interview. Collected data were entered in a predesigned case report form and subsequently analyzed by SPSS 20.

**Results:** The mean age at presentation was 41.59±13.73 (SD) years and most of the cases were male (73%). A total of 90.29% of patients reside in urban areas. Among the positive cases, 13.1% (n = 31) were asymptomatic. Asymptomatic cases were significantly more common in households with 2 to 4 members (p = .008). Both symptomatic and asymptomatic patients shared similar ages of presentation (p = 0.23), gender differences (p = 0.30), and comorbidities (p = 0.11). Only 5.3% of patients received ICU care during their treatment. The most frequent presentation was fever (88.3%), followed by cough (69.9%), chest pain (34.5%), body ache (31.1%), and sore throat (30.1%). Thirty-nine percent (n = 92) of the patients had comorbidities, with diabetes and hypertension being the most frequently observed.

**Conclusion:** There has been an upsurge in COVID-19 cases in Bangladesh. Patients were mostly middle-aged and male. Typical presentations were fever and cough. Maintenance of social distancing and increased testing are required to meet the current public health challenge.

## Background

Coronavirus disease 2019 (COVID-19) emerged in Wuhan, China, in December 2019 and subsequently spread outside China^1^. The virus has spread globally with sustained human to human transmission and affected 216 countries and/or territories with 20, 995, 433 confirmed cases and 760, 774 deaths until 15 August 2020^2^. In Bangladesh, from the first case identified in early March this year, 272,000 individuals tested positive for COVID-19, and 3,591 died^3^. A recent report suggests that the number of COVID-19-related deaths increased by 14.9%, leading to a Case Fatality Rate (CFR) of 1.27%^4^.

The earlier clinical spectrum of COVID-19 was dominated by fever, cough, and shortness of breath^5^. Dyspnea, headache, myalgia, rhinorrhea, sore throat, nausea, and vomiting were also reported in a number of cases^6,7^. In addition to respiratory symptoms, neurological^8^, gastrointestinal^9^, cardiovascular^10^, musculoskeletal^11^, and other systematic presentations have also been reported^6^. It affects all ages, genders, and occupations, especially healthcare workers and family members, with COVID-19 positive cases^3,12–15^. Data up to this day have proven that higher chronological age and presence of comorbidities are the two most common underlying factors responsible for worse outcome^16,17^.

Diagnosis mostly depends on clinical symptoms and signs and subsequent investigations^18^. However, there is increasing evidence that many patients with COVID-19 are asymptomatic or have fewer symptoms to be recognized. The reported prevalence of asymptomatic patients ranged from 1% to 19.2%^19,20^. Unfortunately, asymptomatic cases can transmit the virus to others, consequently acting as a silent harbor of the infection^19^. The previous reports have shown that this virus was isolated from asymptomatic individuals, and the infections had been transmitted from asymptomatic patients^21^. As there are difficulties in screening for asymptomatic infections, it is difficult for national prevention and control of this epidemic unless appropriately addressed. Before that, it is essential to know the detailed picture and differences of both symptomatic and asymptomatic COVID-19 patients’ characteristics and clinical manifestations (if present). As there are limited comparative data on the individuals with symptomatic and asymptomatic COVID-19, this study was conducted to assess the clinico-epidemiological profile of asymptomatic and symptomatic COVID-19-positive patients in Bangladesh. These data would help set effective preventive, and control strategies against this severe public health threat.

## Materials and methods

The first three COVID-19 cases were reported by the Institute of Epidemiology, Disease Control and Research (IEDCR) at 8^th^ March 2020^3^. Since then, the spread of COVID-19 has increased exponentially along with panic. On 26 March 2020, the Government of the People’s Republic of Bangladesh declared general holidays (or lock-down) for 10 days, later extended up to 30 May 2020 to curb the spread of the novel coronavirus in the country^22^. Due to nationwide lock-down, health center-based data collection or community-based national sampling surveys were not feasible. Hence, an online-based cross-sectional survey method was adopted. The study was conducted for the period of 26 April to 27 May 2020. A total of 376 responses were received, and among them, 237 confirmed COVID-19 cases were interviewed. Details of the patient selection are mentioned in **Figure 1**.

**Figure 1.**
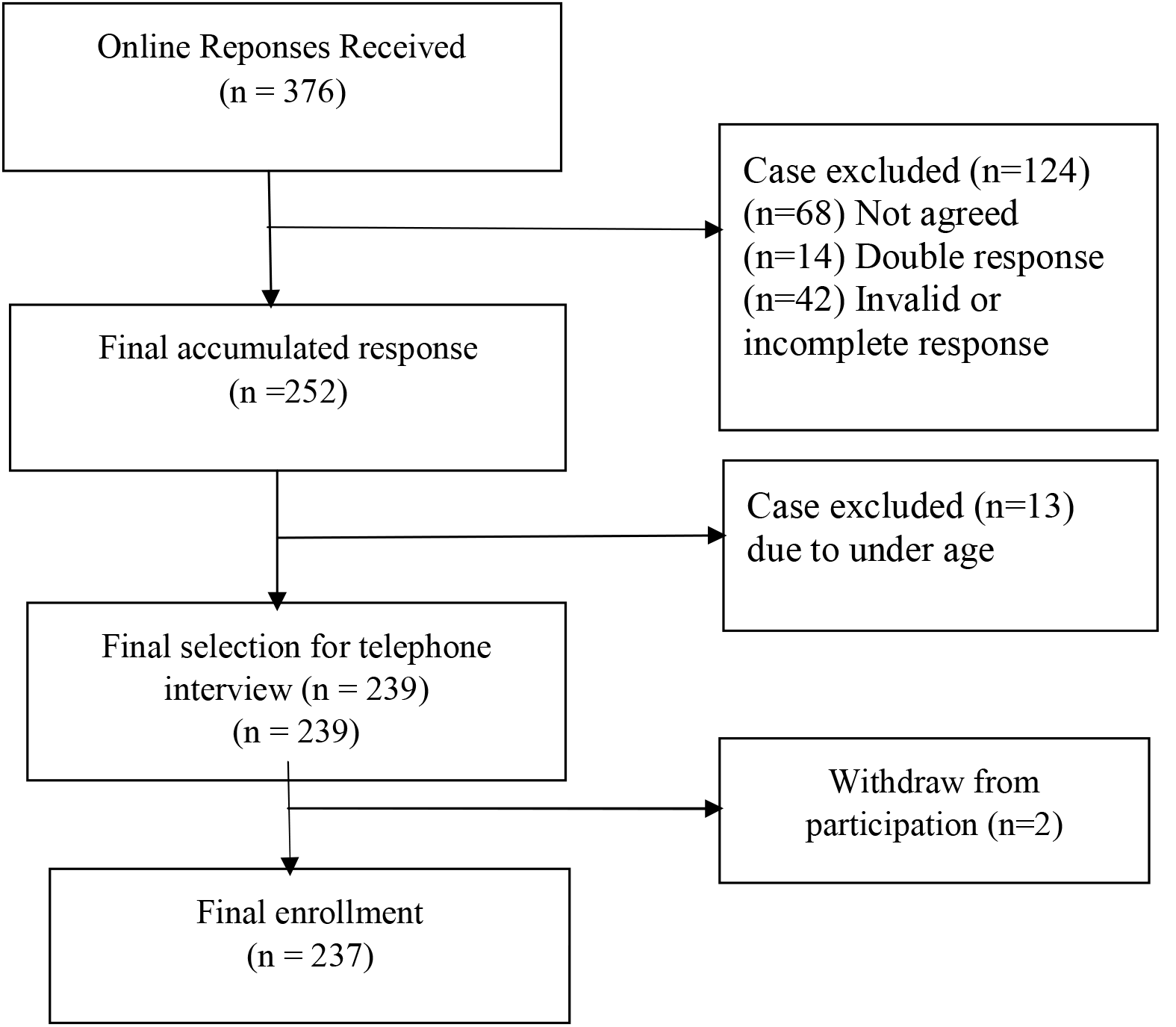
Flow chart of patient selection.

COVID-positive victims aged > 18 years and persons of Bangladeshi nationality were primarily selected as the target group. A recruitment post was posted in the Facebook news feed of the authors. This post contained a brief introduction about the background, objective, procedures, voluntary nature of participation, declarations of anonymity and confidentiality, and notes for filling in the questionnaire and the link to the online questionnaire (Google form). The sensible people of Bangladesh understood the content of the post and agreed to participate in the study were then instructed to complete the questionnaire by clicking the link. After receiving the response and removing duplications, final list of the patients was selected. A telephone-based interview was conducted by three trained physicians mostly to confirm the COVID-19 diagnosis and to reconfirm the data. All patients were requested to provide a snapshot of the test report through messenger or email. Data were collected, emphasizing demographic details and clinical manifestations. Pretesting of the questionnaire (Google form) was performed among ten random participants. The piloting experience was used to make a final adjustment before the final Facebook post was published online. The form was set so that if any person clicked the link to participate and did not agree with the informed consent statements, they could not proceed to the next sections of the questionnaire.

### Ethics statement

Before the commencement of the study, formal ethical approval was obtained from the Ethical Review Committee (ERC) of the Biomedical Research Foundation (BRF), Bangladesh (Memo no: BRF/ERB/2020/003). All methods were performed in accordance with the current Declaration of Helsinki. All participants gave informed consent before participation during completion of the Google form.

### Data cleaning and analyses

Descriptive statistics were used during analysis, where continuous variables were expressed as the mean± standard deviation and categorical variables were expressed as count (percentage). To test the difference, Student’s t-test and chi-square test were performed as appropriate. Data were analyzed with the statistical software SPSS 20 (SPSS Inc., Chicago, IL, USA).

## Results

A total of 237 were finally included in the analysis. Of them, 13.1% (n = 31) were asymptomatic **(Figure 2)**.

**Figure 2:**
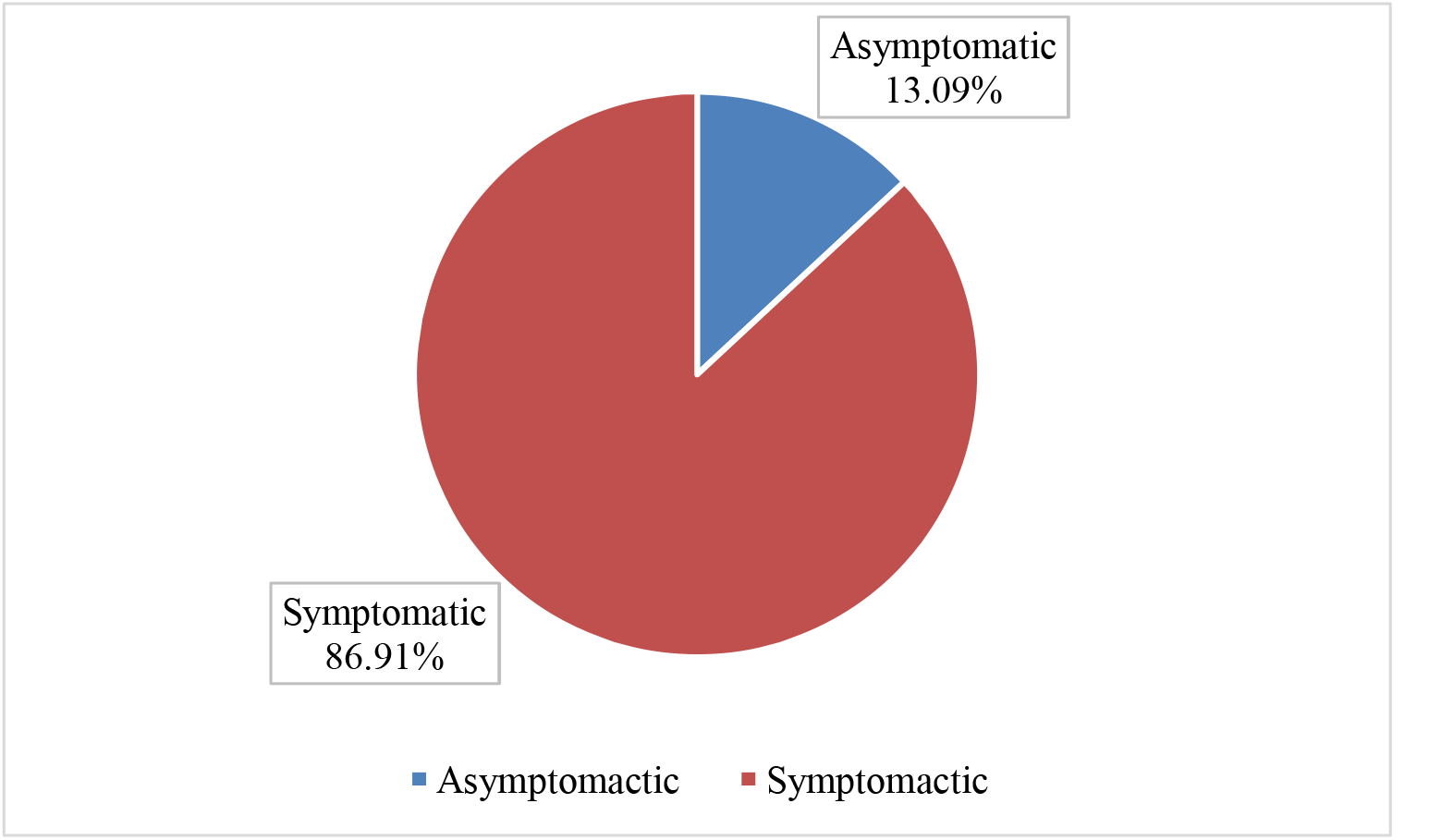
Proportion of asymptomatic and symptomatic and COVID-19 cases.

The mean age of the presentation of asymptomatic cases was slightly lower than that of symptomatic cases (39 vs. 41 years; p = .26). Both males and females are equally affected by the disease (p = .48) and predominately resided in urban areas. Asymptomatic cases were mostly observed in the house where 2–4 people lived within the same household. Twenty-six percent of the asymptomatic and forty-one percent of symptomatic cases had at least one comorbid disease. Among them, diabetes mellitus and hypertension were most frequently observed in both groups. More details about the socio-demographic profile are shown in **table 1**.

**Table 1.**
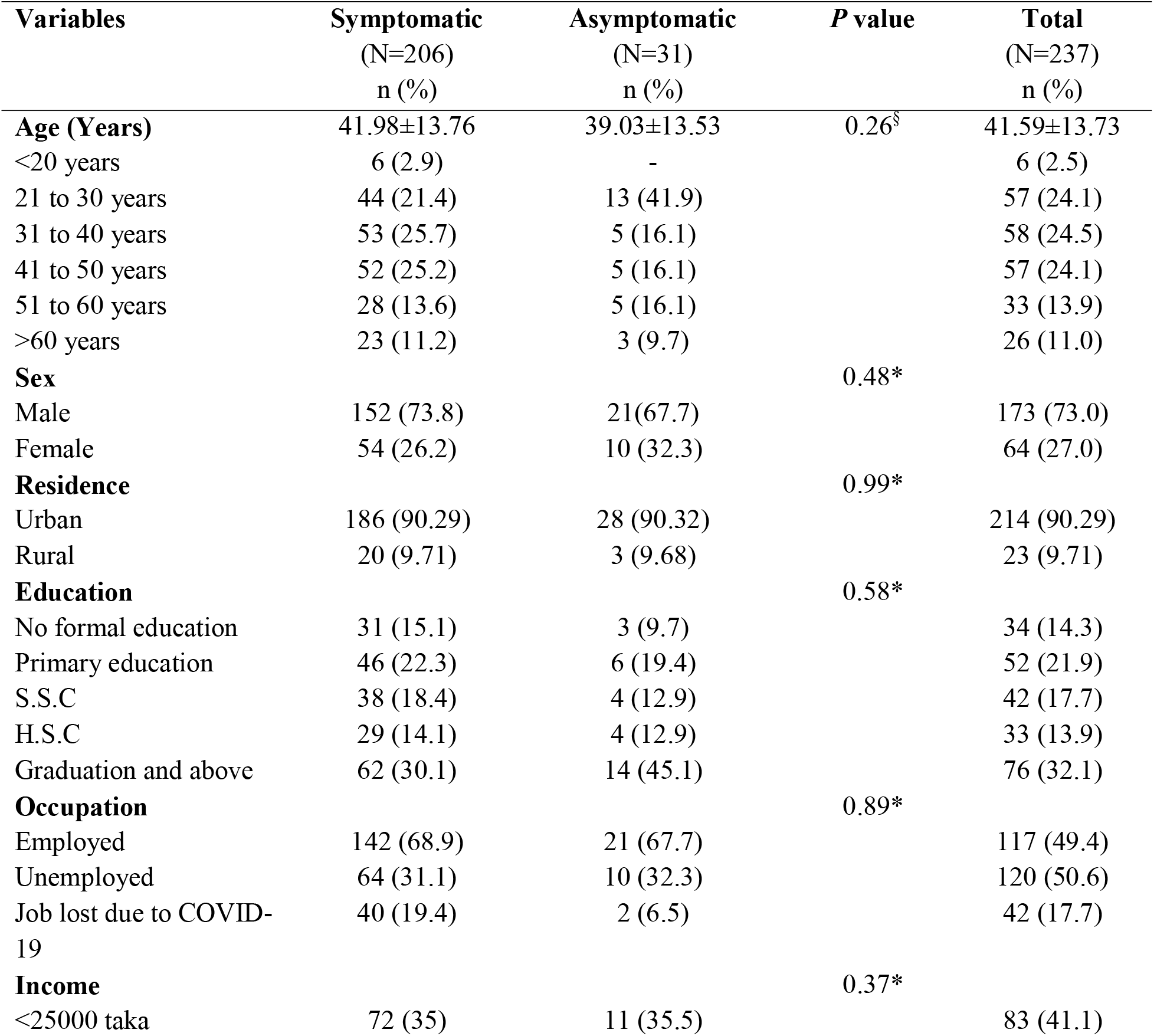
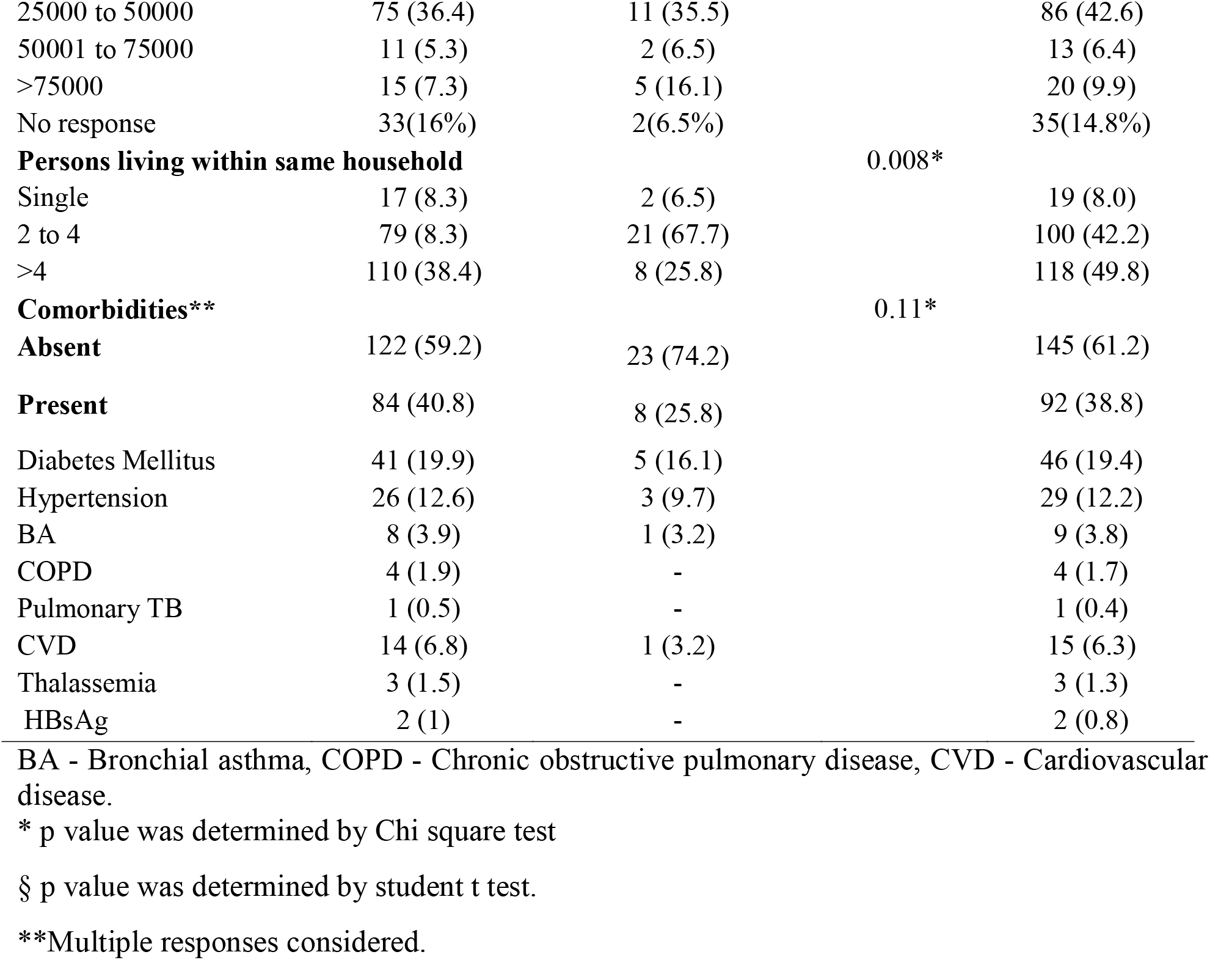
Characteristics of person with COVID-19.

Among symptomatic COVID-19 patients, the average duration of illness was nine days, ranging from 6 to 21 days. Fever was the most common symptom (88.3%;182/206), followed by cough (69.9%; 144/206) and chest pain (34.5%, 71/206). Intensive care support was warranted in 5.3% of cases (11/206). The detailed clinical characteristics of symptomatic patients are shown in **Table 2**.

**Table 2.**
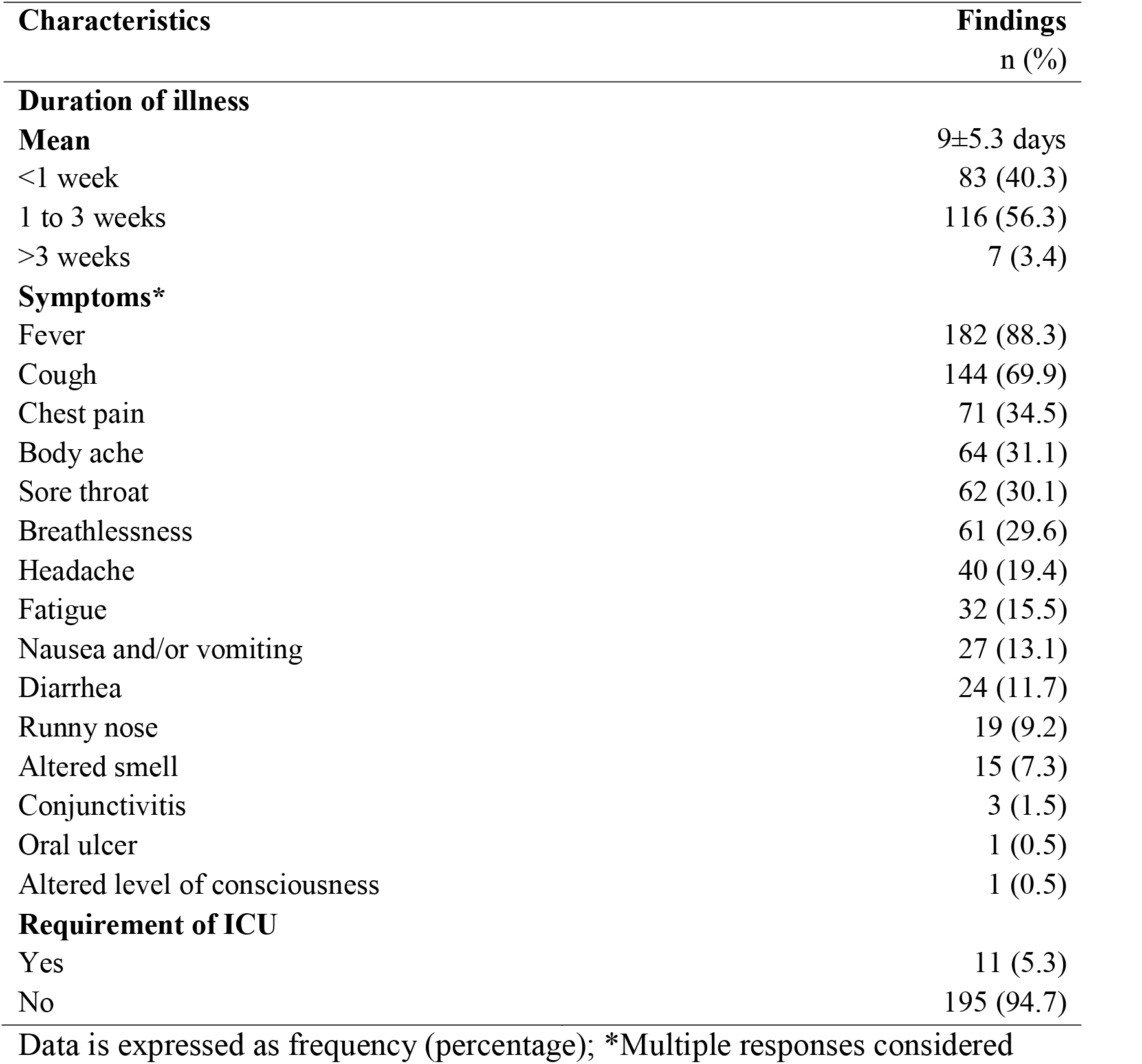
Clinical characteristics of symptomatic patients (n = 206)

| **Characteristics** | **Findings**<br>**n (%)** |
| --- | --- |
| **Duration of illness** |  |
| **Mean** | 9±5.3 days |
| <1 week | 83 (40.3) |
| 1 to 3 weeks | 116 (56.3) |
| >3 weeks | 7 (3.4) |
| **Symptoms\*** |  |
| Fever | 182 (88.3) |
| Cough | 144 (69.9) |
| Chest pain | 71 (34.5) |
| Body ache | 64 (31.1) |
| Sore throat | 62 (30.1) |
| Breathlessness | 61 (29.6) |
| Headache | 40 (19.4) |
| Fatigue | 32 (15.5) |
| Nausea and/or vomiting | 27 (13.1) |
| Diarrhea | 24 (11.7) |
| Runny nose | 19 (9.2) |
| Altered smell | 15 (7.3) |
| Conjunctivitis | 3 (1.5) |
| Oral ulcer | 1 (0.5) |
| Altered level of consciousness | 1 (0.5) |
| **Requirement of ICU** |  |
| Yes | 11 (5.3) |
| No | 195 (94.7) |

Data is expressed as frequency (percentage); \*Multiple responses considered

## Discussion

Severe acute respiratory syndrome coronavirus 2 (SARS-CoV-2) infection emerged in China in December 2019 and was announced a Public Health Emergency of International Concern (PHEIC) by the WHO and became a pandemic around the world. The complete clinical manifestation is not clear to date, as the reported symptoms range from mild to severe, even asymptomatic, in many cases with a wide-ranging variety^19,21,23–25^. A number of evidences have shown that asymptomatic patients can effectively spread the virus, and the emergence of these silent spreaders of SARS-CoV-2 has caused difficulties in the control of the pandemic^21,23^. Detailed information on the prevalence of asymptomatic cases of coronavirus disease 2019 (COVID-19) and the clinical characteristics of mild COVID-19 is essential for effective control of the COVID-19 pandemic. Therefore, this study aimed to report the clinico-epidemic profile of asymptomatic and symptomatic COVID-19 patients in Bangladesh.

In our study, we found that 13.09% of COVID-19 patients were asymptomatic. Kim et al. found 41 (19.2%) individuals out of 213 cases with COVID-19 were asymptomatic until admission^19^. Data from Long et al. showed that 20.8% of patients with COVID-19 had asymptomatic infections^24^. Li et al. found a relatively higher prevalence (29.4%) of asymptomatic patients^26^, consistent with another study that observed that 34.6% of COVID-19 patients were asymptomatic onboard the Princess Cruise^27^. Considering the different models for calculating the estimated proportion of asymptomatic patients, a meta-analysis by Kronbichler et al. found that asymptomatic cases could account for more than 20% of all COVID-19 patients^28^. However, our findings might not be an accurate approximation of the asymptomatic individuals in the general population, as asymptomatic infections were identified from those who were at high risk for infection (including close contacts or had a history of travel to hotspot zones) and not from a random sample of people. Therefore, the prevalence of asymptomatic infections needs to be evaluated through population screening^29^. Additionally, the proportion of asymptomatic infections might be even higher, as some cases might be missed by RT–PCR testing. Long et al. found seven asymptomatic patients with COVID-19 infection with negative RT–PCR results^30^.

We found an overall male (73%) predominance without any significant difference between symptomatic and asymptomatic groups (p>0.05). However, previous studies found that asymptomatic infection is not limited to young or middle-aged adults^31^ but also infants^31^, children^32^, and even the elderly^33^. In addition, asymptomatic infections were found in both males^34^ and females^21^. Interestingly, in our study, asymptomatic cases were significantly higher compared to symptomatic cases in the house where 2–4 people lived within the same household (67.7% vs. 8.3%, p = .008). No other evidence is available to explain the findings and needs to be investigated further.

Among the symptomatic cases, the majority had one or more symptoms. The most prevalent presentation was fever followed in decreasing order by cough, chest pain, body ache, sore throat, and breathlessness. These pattern of symptoms are consistent with the findings of other studies^35,36,37^. The initial studies in China^35,36,38,39^ also found predominant presentation of fever and cough among COVID-19 patients, which appears to be more common in adults than children, as well as breathlessness, and body ache, among other clinical features. A meta-analysis of Rodriguez et al found a similar frequency of fever in COVID-19 with SARS and MERS, but the cough frequency was higher in COVID-19 than MERS (< 50%)^36^

We found a lower proportion of symptomatic patients with complaints about hyposmia (7.3%) compared to other studies where the proportion are relatively large^19,38,40^. These could be explained by the underreporting status of the patients or all of the patients may not be aware about the symptoms, or due to stuffy nose or runny nose patients may assume that it is usual consequences of stuffy nose rather a specific feature of COVID-19. Therefore, further studies are needed to determine the accuracy of this symptom in diagnosing COVID-19 and to determine whether this distinctive symptom is associated with nasal congestion or dysfunction of the olfactory nerve due to COVID-19 or other comorbidities^19^. However, headache, fatigue, nausea and/or vomiting, diarrhea, runny nose, stuffy nose, conjunctivitis, and oral ulcer were also found in small proportions in this study. Nevertheless, we should not underestimate these symptoms for the diagnostic purpose of COVID-19.

This study has several limitations. There was an oversampling of a particular network of peers (e.g., socially engaged), leading to selection bias. As a result, the conclusion is less generalizable to the entire population, particularly for less educated people. More detailed patient information, particularly clinical outcomes and detailed investigations, was unavailable. The scope of confirmation of the data regarding comorbid conditions was minimal. Therefore, our findings on the symptoms of individuals with COVID-19 should be considered with caution.

## Conclusion

In conclusion, much attention should be given to identifying asymptomatic patients to reduce the transmission of SARS-CoV-2, as their potential to spread the virus cannot be misjudged. If both asymptomatic and symptomatic COVID-19 patients can be appropriately quarantined, community and family clustered transmission may be greatly reduced. Contact tracing and testing of the suspected case could be another strategy for preventing and controlling the disease. We have summarized the clinical and epidemic characteristics of both symptomatic and asymptomatic patients with COVID-19 with severe limitations. This deserves further study in outpatient, primary care, or community settings to obtain a full picture of the spectrum of the clinical scenario.

### Future scope and recommendation of the study

The data in this study permit an early assessment of the epidemiological and clinical characteristics of Bangladesh’s COVID19 cases. Furthermore, these data could be used in future planning and implementation of prevention and control strategies.

## Data Availability

Data will be available on request

## Declarations

## Supplementary Materials

Available on request

## Author Contributions

Conception and development of the idea: MJH

Writing – Original Draft Preparation: MJH, JF, MR, SMC,

Data analysis: SMC

Data collection: SR, JF, MR

Writing – Review & Editing: MJA, MASK, MR, MTH, ER, MRA

## Funding

This research received no external funding.

## Acknowledgments

The author thanks the patients who participated in the study during this stressful situation. The authors are also thankful to the research team of the ‘Pi Research Consultancy Center’ http://www.pircc.org) for support throughout the study period, especially for journal selection and manuscript formatting. The team also acknowledges the role of Dr Bahaduddin, Dr Jobayer Amin and Dr Muhibur Rahman Jewel for their cordial support during the support and inspiration.

## Conflict of Interests

The authors declare that there are no conflicts of interest regarding the publication of this paper.

## Ethical consideration

Ethical measures were taken throughout the study period to maintain a high standard of confidentiality and anonymity of the participants. Formal ethical clearance was taken from the ethical review committee of the Biomedical Research Foundation (BRF) for conducting the study, and formal permission was taken from the responders through Google form.

## Consent of Publication

All authors agree to publish the article.

## List of abbreviations

BRF: Biomedical Research Foundation
CFR: Case Fatality Rate
COVID-19: Corona Virus Disease 2019
ERC: Ethical Review Committee
GAD: Generalized Anxiety Disorder
ICU: Intensive Care Unit
IEDCR: Institute of Epidemiology, Disease Control and Research
PHEIC: Public Health Emergency of International Concern
PHQ: Patient Health Questionnaire
SARS: Severe Acute Respiratory Syndrome
SPSS: Statistical Software for Social Science
SSC: Secondary School Certificate

## Notes

### Competing Interest Statement

The authors have declared no competing interest.

### Funding Statement

No fund received.

### Author Declarations

Before the commencement of the study, formal ethical approval was obtained from the Ethical Review Committee (ERC) of the Biomedical Research Foundation (BRF), Bangladesh (Memo no: BRF/ERB/2020/003).

